# *Staphylococcus lugdunensis* and Coagulase-Negative Staphylococci species characterisation in a tropical climate

**DOI:** 10.1101/2022.10.12.22280773

**Authors:** Matthew Howes, Rob Baird

## Abstract

Coagulase negative staphylococci (CoNS) are a heterogeneous group of colonising organisms that have historically been differentiated from Staphyloccocus aureus but are now known to cause invasive disease. We characterised the CoNS species from a single tropical centre to compare with previous studies. 1262 isolates were collected with 203 from blood stream samples over an 18 month period. S. lugdunensis was the second most isolated CoNS comprising 15.9% of isolates, much greater than comparative studies. A single blood stream infection with S. lugdunensis was identified over this time period, comprising a much lower ratio of blood to swab isolates than in previous studies.

## Introduction

Coagulase negative staphylococci (CoNS), a heterogeneous group of human and animal colonisers, historically were important to differentiate from their virulent relative, *Staphylococcus aureus*, but now many are recognised as causing invasive disease. Biochemical and genomic speciation has become more widely and routinely available, allowing the CoNS species distribution in human clinical samples to be better defined (1). We characterised the CoNS species from a single tropical tertiary centre for both swab and blood isolates, and review and compare with previous studies (2). This study was ethics approved: EFILE2022/25273.

## Methods

Geographically, Royal Darwin Hospital is the tertiary referral hospital for the Top End of the Northern Territory, a region of over 500,000 km^2^ with a tropical climate (latitude 12.5 *S). All clinically isolates of CONS were speciated from 1^st^ January 2019 until 30^th^ June 2022 were included, with duplicate patient specimens excluded. During this time there were also 7546 *S. aureus* isolates (including 161 [2.1%] from bloodstream isolates). Isolates were biochemically identified by Vitek 2 (Biomerieux, St Louis USA) and characterised as blood or swab isolates. There were 1262 isolates collected, of which 203 (16%) were from blood stream samples. The results are presented in Table 1.

## Results

*S. epidermidis* was predominant, 30.8% of all CONS isolates, and over 50% of the blood stream isolates; *S. saprophyticus* predominantly in urine, and *S. epidermidis* like organisms (*S. haemolyticus, S. capitis* and *S. hominis*) were also frequent in swab and blood stream isolates.

*S. lugdunensis* was the second most commonly isolated CONS species identified with 201 isolates, 15.9% of all isolates. This frequency is far higher than other published results (3). 24% and 66.3% of these isolates came from the pelvic and lower body sites respectively, as described previously (4).

However, despite the large number of skin swab isolates, there was only a single blood stream isolate of *S. lugdunensis* detected during this study from a patient admitted with fever and found to have *S. lugdunensis* in one blood culture bottle only. Repeat blood cultures were negative three days later. The patient had a presumed ileostomy source and received 2 weeks of intravenous antibiotics for bacteraemia, and the patient remained well on review.

## Discussion

Though 15.9% of all CONS isolates, *S. lugdunensis* represented only 0.5% of CONS blood stream isolates, with a low 0.5% rate of blood to swab isolates. Our rate of blood-stream *S. lugdunensis* isolation was much lower than comparable sized series (1). A further retrospective look back over 10 years identified only two other blood stream isolates, both one bottle only, and both not deemed clinically significant. This low incidence of serious blood stream infection over 10 years has not been reported previously form tropical settings (5).

We plan to sequence a number of local *S. lugdunensis* isolates to determine the presence of postulated virulence factors, and compare phylogenetic relationships with known virulent strains, to characterise this epidemiological observation.

## Data Availability

Data is stored on a secure server. Non-confidential anonymised data is available on request from the authors.

