## Supplementary material for "*Staphylococcus lugdunensis* and Coagulase-Negative Staphylococci species characterisation in a tropical climate": Table 1

Table 1. Characterisation of swab and blood stream isolates of CoNS between 2019 and 2022 at Royal Darwin Hospital,Northern Territory, Australia

| **Species** | **All sites isolates number and frequency N** (%) | **Blood isolates N** (% of total CoNS) | **% blood versus all sites isolates** |
|---|---|---|---|
| *S. epidermidis* | 389 (30.8) | 104 (51.2) | 26.7% |
| *S. lugdunensis* | 201 (15.9) | 1 (0.5) | 0.5% |
| *S. saprophyticus** | 193 (15.3) | 3 (1.5) | 1.6% |
| *S. haemolyticus* | 184 (14.6) | 28 (13.8) | 15.2% |
| *S. capitis* | 55 (4.4) | 23 (11.3) | 41.8% |
| *S. hom.hominis* | 44 (3.5) | 23 (11.3) | 52.3% |
| *S. caprae* | 38 (3.0) | 4 (2.0) | 10.5% |
| *S. simulans* | 36 (2.9) | 1 (0.5) | 2.8% |
| *S. lentus* | 29 (2.3) | 4 (3) | 13.8% |
| *S. pseudintermedius* | 24 (1.9) | 2 (1.0) | 8.3% |
| *S. sciuri* | 21 (1.7) | 1 (0.5) | 4.8% |
| *S. warneri* | 14 (1.1) | 2 (1.0) | 14.3% |
| *S. auricularis* | 13 (1.0) | 4 (2.0) | 30.8% |
| *S. cohnii cohnii* | 10 (0.8) | 2 (1.0) | 20.0% |
| *S. xylosus* | 4 (0.3) | 0 (0) | 0.0% |
| *S. carn.carnosus* | 3 (0.2) | 0 (0) | 0.0% |
| *S. schleiferi* | 2 (0.2) | 0 (0) | 0.0% |
| *S. gallinarum* | 1 (0.1) | 0 (0) | 0.0% |
| *S. kloosii* | 1 (0.1) | 1 (0.5) | 100.0% |
| **Total** | **1262** (100) | **203** (16.1) | |

* predominantly urine isolates
